# Integrated Palliative Care in Rural Communities: A Qualitative Study of Service Integration in Southern Minnesota

**DOI:** 10.64898/2026.03.25.26349325

**Authors:** Meritxell Mondejar-Pont, Ellen Vorbeck, Kristen Abbott-Anderson

**Author notes:** **Corresponding Author:** Meritxell Mondejar-Pont.

## Abstract

**Background:** Palliative care services improve quality of life and health outcomes for individuals living with chronic and life-limiting illnesses. Although these services have expanded considerably in urban areas, their availability remains limited in many rural communities. This study aimed to identify key components of integrated palliative care services and examine how these elements are implemented within rural healthcare systems in southern Minnesota.

**Methods:** A qualitative case study using deductive content analysis was conducted. Semi-structured interviews were carried out with healthcare professionals involved in palliative and hospice care serving rural communities in southern Minnesota.

**Results:** Participants identified several essential components of integrated palliative care, including multidisciplinary care teams, continuity of care across healthcare settings, interprofessional collaboration, and early identification of patients who may benefit from palliative care. Existing services in southern Minnesota incorporate several integrated elements, such as coordinated care teams, individualized care plans, nurse-led case management, professional training, and the use of virtual visits for geographically distant patients. However, participants also identified important gaps, including limited availability of palliative care services in rural areas, fragmented continuity of care, challenges in early patient identification, funding and insurance barriers, and the absence of a unified palliative care network.

**Conclusions:** While palliative care services in southern Minnesota demonstrate important strengths, further efforts are required to improve service integration, coordination, and access for rural populations. Strengthening integrated PCSs may help reduce disparities in access to care and improve service delivery for rural patients and their families. These findings may inform the development of integrated palliative care models in rural healthcare systems beyond the study setting.

## 1. BACKGROUND

Since its recognition as a medical specialty in 2006, palliative care services (PCS) have been widely documented as significantly improving health outcomes for individuals living with chronic and life-threatening illnesses(1–3). Evidence suggests that integrated palliative care services (IPCS) can reduce symptom burden, decrease healthcare utilization and associated costs, promote health literacy, facilitate goals-of-care discussions, and provide psychosocial support for patients, families, and caregivers (2,4–6). As a result, PCS have experienced substantial growth in metropolitan and urban areas of the United States. However, this expansion has not occurred at the same rate in rural communities.

Rural areas, defined as non-metropolitan counties with populations living in towns and surrounding areas of up to 50,000 residents (7), remain underserved in terms of access to PCS. Limited growth of PCS in these regions has contributed to disparities in care availability and access. These challenges provide the rationale for examining PCS in southern Minnesota(8–12).

The purpose of this qualitative case study was to identify perceived key components of ideal PCS, including integrated elements that function effectively and those requiring improvement for rural populations in southern Minnesota. Previous research has identified several facilitators for the successful implementation of IPCS, including clear definitions of palliative care, timely referrals, specialized training for healthcare providers, consistent policies and funding, and coordination of services across micro, meso, and macro levels of healthcare systems (8–10,12–14). When these facilitators are absent, they can become barriers that further complicate the provision of IPCS in underserved rural areas.

For the purposes of this study, PCS are defined according to the WHO (WHO). Palliative care is an approach that seeks to improve the quality of life of adults, children, and their families facing problems associated with life-threatening illness. This approach focuses on preventing and alleviating suffering through early identification, comprehensive assessment, and treatment of pain and other physical, psychosocial, and spiritual problems(15).

IPCS align with this definition and incorporate the National Consensus Project Clinical Practice Guidelines for Quality Palliative Care(16). These guidelines outline key domains, including care structure and processes, physical and psychological care, social determinants of health, spiritual and cultural considerations, ethical and legal aspects, and care at the end of life. However, the interpretation and implementation of these domains may vary across healthcare systems, making consensus regarding the delivery of IPCS challenging.

Despite growing evidence on palliative care delivery, limited research has examined how integrated PCS are structured and experienced within rural healthcare systems in the United States.

This study, therefore explored facilitators and barriers to the provision of integrated PCS and examined their alignment with existing literature. Semi-structured interviews were conducted with palliative and hospice healthcare providers serving rural communities in Blue Earth, Brown, and Nicollet counties in southern Minnesota.

## 2. MATERIALS AND METHODS

This study aimed to replicate a study previously conducted in a rural region of Spain to evaluate the level of integration of PCS provided in southern Minnesota. This study followed the COREQ (Consolidated Criteria for Reporting Qualitative Research) guidelines for qualitative research reporting (17). The COREQ checklist is provided as Supplementary Material.

To achieve this objective, a qualitative case study design was employed, following a similar methodological approach to that described in research conducted in a rural Spanish community (18). The study focused on examining PCS in three rural counties of southern Minnesota: Blue Earth, Brown, and Nicollet.

The specific objectives of the study were: 1) to identify the key elements of an ideal IPCS, and 2) to determine which elements are currently present or missing within the existing PCS and how these influence the level of service integration.

Following approval from the institutional review board, participant recruitment was conducted between June 1st and August 30, 2022, among palliative care and hospice healthcare professionals working in Blue Earth, Brown, and Nicollet counties. The research team identified individuals working in administrative or direct care roles in order to include a diverse range of professionals, including physicians, nurses, managers, and allied healthcare professionals.

Initial contact was made by a member of the research team with individuals holding administrative roles within palliative care or hospice organizations. These contacts were asked to identify additional professionals involved in administrative or clinical roles within PCS. Potential participants were then asked to provide permission for the researchers to contact them.

Once permission was obtained, typically through email, the research team sent a recruitment email describing the study. The email included a link to a written informed consent form, which participants were required to review and sign before participation. Written informed consent was obtained electronically from all participants, and signed consent forms were securely retained by the research team for record-keeping purposes. Participants were then able to indicate their preferred day, time, and format for the interview.

Seven participants volunteered to participate in the study, including two palliative care directors, one palliative care manager, one administrator/lead physician, two Doctor of Nursing Practice (DNP) and Advanced Practice Registered Nurses (APRNs), one APRN, and one physician assistant. Participants were given the option to meet with the researcher prior to the interview to learn more about the study and ask questions. The sample size is consistent with qualitative case study research, where smaller samples are often used to obtain in-depth perspectives from participants with relevant expertise. Recruitment continued until thematic saturation was reached, meaning that no substantially new information or perspectives were emerging from additional interviews.

The study involved a one-time semi-structured interview, that lasted approximately 60 minutes, conducted by members of the research team. The researchers conducting the interviews had prior experience in qualitative research and palliative care services research. Participants were asked to describe what would constitute an ideal palliative care service and how the current PCS in their region aligns with that ideal system. This approach allowed researchers to explore similarities and gaps between ideal and existing services (see Appendix 1 for the interview guide).

Interviews were audio and/or video-recorded with participant permission and conducted using the Zoom platform. No non-participants were present during the interviews. Following the interviews, transcripts were reviewed and cleaned by the researcher who conducted the interview to ensure accuracy and remove identifying information.

Two members of the research team independently analysed the interview transcripts using the analytical framework developed in the previously mentioned study (18) and following the model of directed content analysis described by (19). Initial codes were generated from the predefined framework and applied to the interview transcripts. These preliminary codes were then grouped into broader categories and compared across cases. The coding framework used in the directed content analysis, including the main categories, preliminary codes, grouped codes, and generic categories, is presented in Table 1.

**Table 1.** Analytical framework including main categories, preliminary codes and generic categories.

| MAIN CATEGORIES | PRELIMINARY CODES | GROUPS OF CODES | GENERIC CATEGORIES |
| --- | --- | --- | --- |
| Multidisciplinary /<br>Interdisciplinary<br>teams | PC specialists | Integrated<br>multidisciplinary and<br>interdisciplinary teams | Comprehensive team of<br>palliative care professionals |
| Case management | Nurse | Nurse coordinator | Nurse-led care coordination |
| Coordination | Coordination, collaboration | Team coordination and<br>collaboration | Coordination within<br>healthcare services |
| Continuity of care | Integrated approach and<br>interconnection of services<br>(inpatient, outpatient,<br>hospital, primary care, home<br>care, community care) | Integrated approach<br>connecting inpatient,<br>outpatient and<br>community services | Continuity of care across<br>healthcare levels |
| Early intervention of IPC | Early identification of patients; avoiding PC provision only in the last six months of life | Early referral and integration of PC services | Early identification and provision of palliative care to patients in need |
| Patient-centred care | Holistic care; individualized care plans; advance care directives; family involvement | Holistic patient-centred care with individualized care plans | Holistic and individualized care for patients and families |
| Training and education | Need for healthcare professionals' PC training; family education on PC | Education for professionals and families | Education and training in palliative care |
| Supportive policies and leadership | Need for PC policies | Supportive policy framework | Supportive palliative care policies |
| Shared information systems | Electronic health records; shared communication systems | Electronic shared communication systems | Shared information systems for patient records and professional communication |
| Community-based palliative care | Outpatient and community PC services | Community-based PC services | Provision of outpatient/community PCS |
| Support for grieving processes | Bereavement support | Bereavement services | Provision of bereavement support services |
| PC system organization | Need to better organize PC services and connect them with community services | Coordinated PC network | Organized PC network connecting healthcare and community services |
| Patient symptom management | Pain control and symptom management | Improved symptom control | Improved pain and symptom management |
| Virtual visits (rural patients) | Virtual care and face-to-face care | Hybrid care delivery | Provision of virtual and face-to-face visits for palliative care patients |

After independent coding, the two researchers met to discuss the findings and resolve discrepancies. When disagreements occurred, they were reviewed with a third researcher until consensus was reached. This process of independent coding and consensus discussions helped enhance the credibility and trustworthiness of the qualitative analysis.

## 3. RESULTS

The findings of this study addressed two primary objectives: 1) to identify the integrated elements that should be present in an ideal IPCS, and 2) to examine which of these elements are currently present within the Southern Minnesota Palliative Care Services (SMPCS) and which require improvement.

### 3.1 Essential Elements for an Ideal Integrated Palliative Care System

The key themes and subthemes identified through deductive analysis are summarized in Table 2, which presents the essential elements of an optimal integrated palliative care system (IPCS). The main elements identified through the interviews are described below.

**Table 2.**
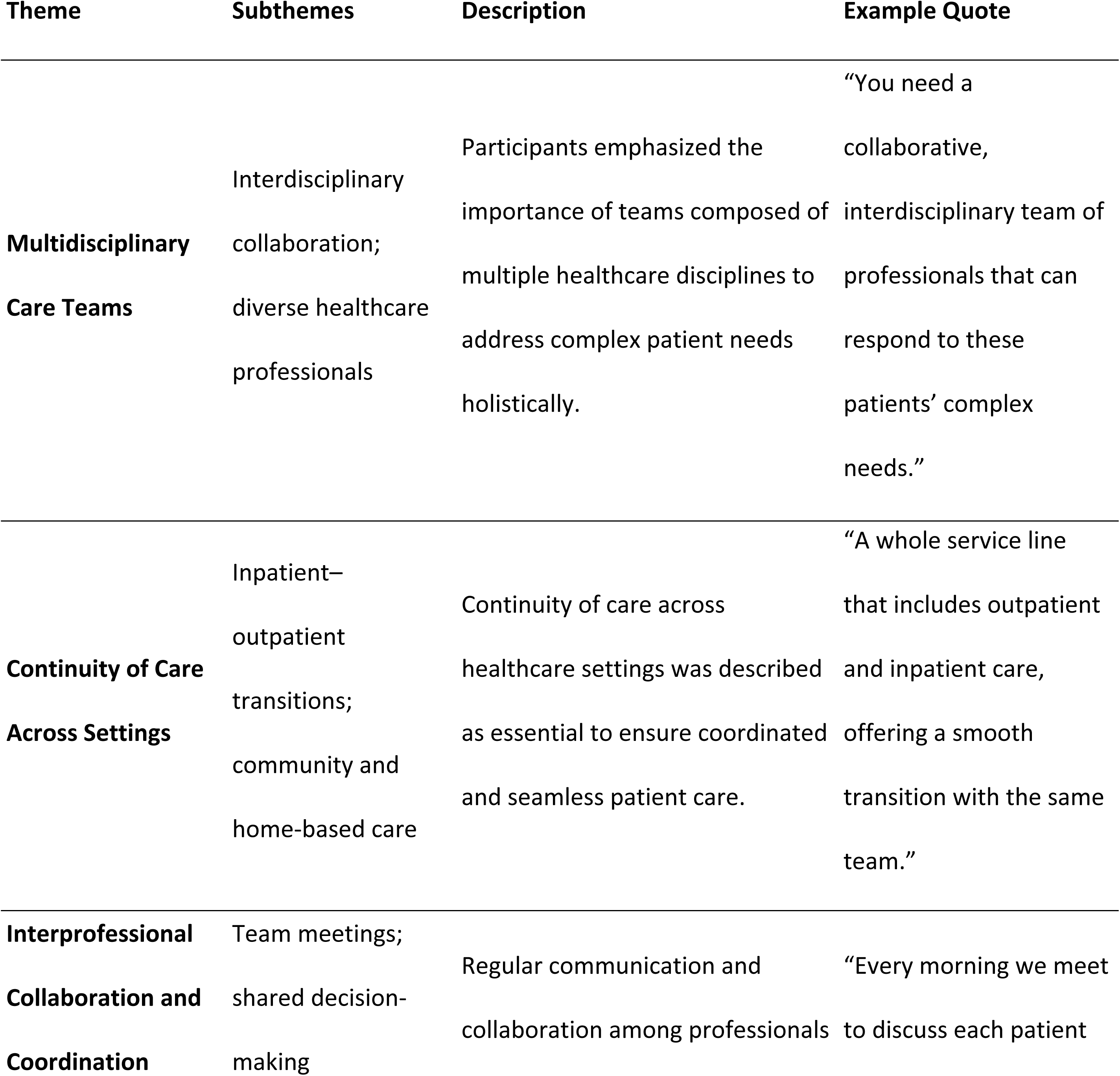
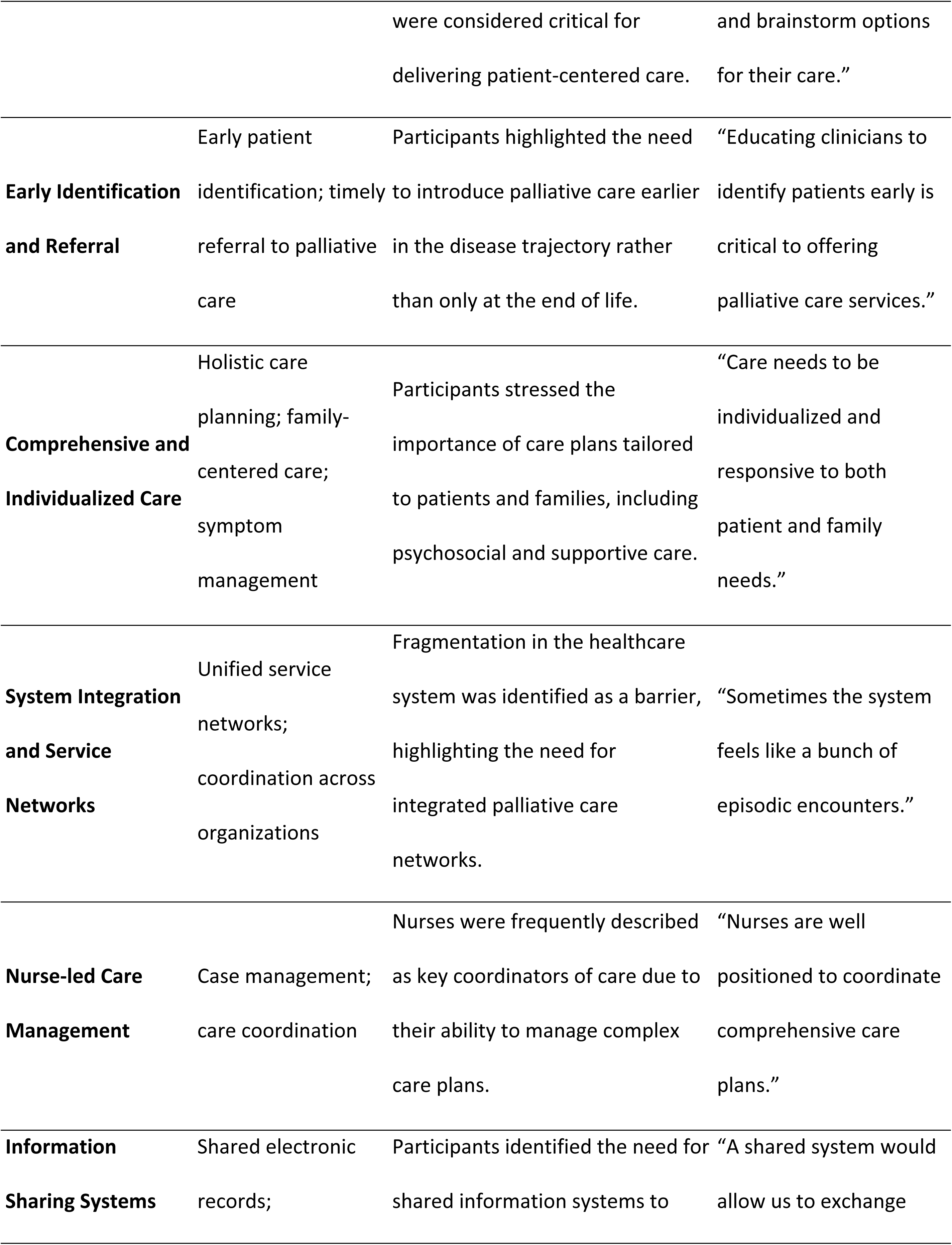
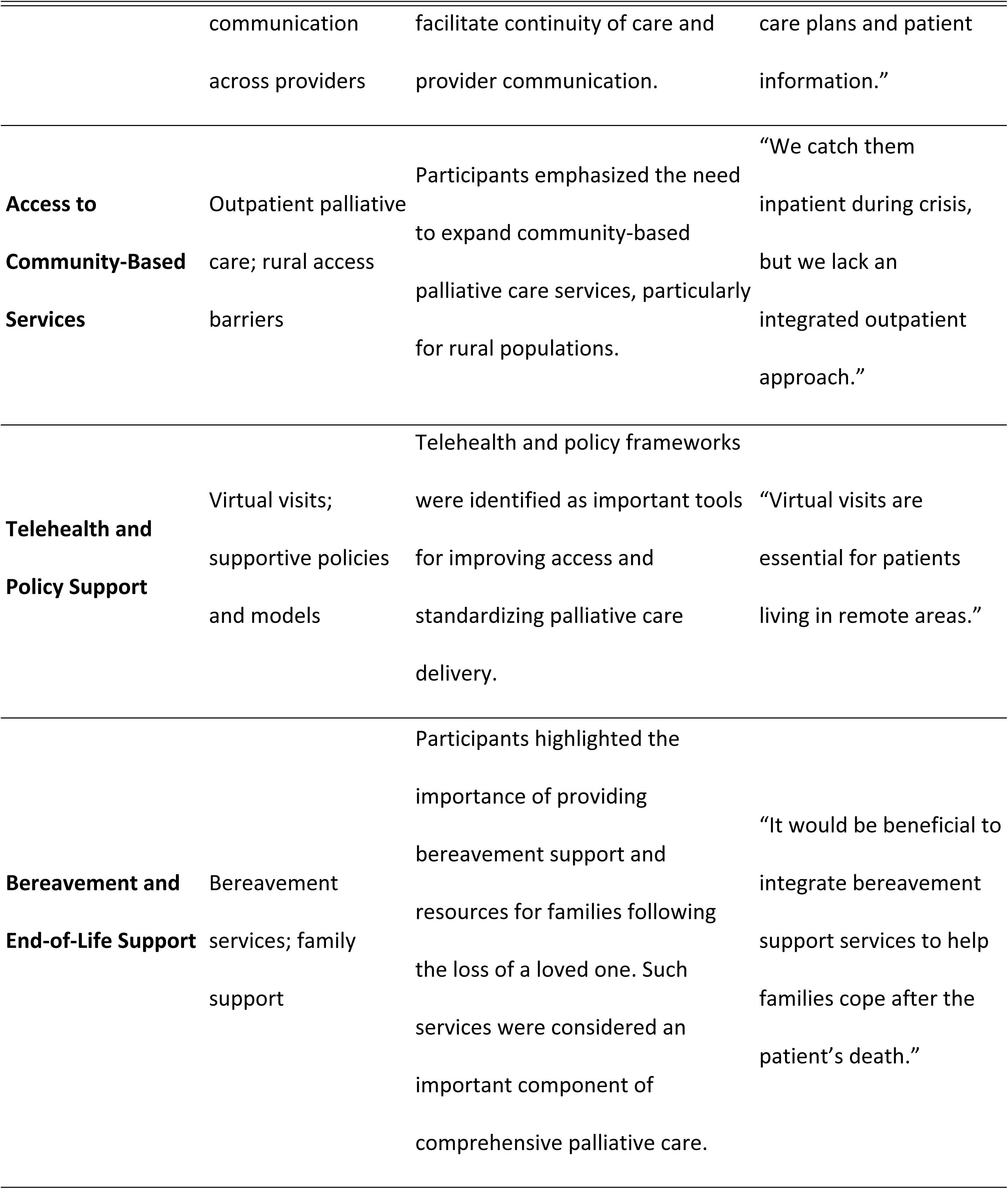
Key Themes and Subthemes Identified for Integrated Palliative Care Services.

**Multidisciplinary teams** were consistently identified as a fundamental component of effective PCS. Participants emphasized the need for collaborative, interdisciplinary teams capable of addressing patients’ complex needs through a holistic approach to care. As one participant explained, *“you need a collaborative, interdisciplinary team of professionals that can respond to these patients’ complex needs and provide robust, holistic palliative care.”*

Another frequently mentioned element was **continuity of care across service levels**. Participants highlighted the importance of ensuring coordinated transitions between inpatient care, outpatient services, primary care, home care, and community-based services. One interviewee described this as *“a whole service line that includes outpatient and inpatient care, offering a smooth transition with the same team.”*

**Interprofessional collaboration and coordination** were also emphasized as essential components of IPCS. Participants described how regular team meetings and collaborative decision-making processes support effective patient-centered care. One participant noted that *“every morning we meet to discuss each patient, which helps us brainstorm the best options to meet their needs.”*

Approximately half of the participants emphasized the importance of **early identification of patients who could benefit from palliative care**. Participants suggested that improved education for healthcare providers could facilitate earlier referrals and prevent PCS from being introduced only during the final stages of illness.

Participants also highlighted the importance of **comprehensive and individualized care** that addresses both patient and family needs throughout the disease trajectory. This includes symptom management, goals-of-care discussions, advance care planning, and bereavement support.

Another key element identified was the need for **well-organized palliative care networks** to improve coordination and access to services. Participants noted that the current healthcare system can be fragmented, which may limit timely access to palliative care in rural communities.

Several participants also emphasized the importance of **nurse-led care management**, noting that nurses are particularly well positioned to conduct comprehensive assessments and coordinate individualized care plans. In addition, participants highlighted the importance of **shared information systems** to facilitate communication between providers and ensure continuity of care through access to electronic patient records and care plans.

Participants also identified several additional elements that could strengthen integrated palliative care systems (IPCS), including the expansion of **community-based and outpatient PCS**, the use of **telehealth or virtual visits** to reach patients living in remote areas, and the development of **supportive policies** and standardized implementation models. These elements were described as important mechanisms for improving access to services and facilitating the coordination of care within rural healthcare systems.

Participants further emphasized the importance of providing **bereavement support for families** following the death of patients receiving palliative care. Bereavement services were described as an essential component of comprehensive palliative care, helping families cope with loss and ensuring continuity of support beyond the patient’s death. Several participants noted that integrating bereavement resources within palliative care programs could strengthen family support during the end-of-life process and improve the overall continuity of care.

### 3.2 Elements in Southern Minnesota Palliative Care Services Requiring Improvement

Participants identified several strengths within the existing SMPCS, as well as areas requiring further development to achieve greater system integration.

A comparison between the ideal elements of an integrated palliative care system and the current situation in SMPCS is presented in Table 3.

**Table 3.** Comparison Between Ideal IPCS Elements and Current SMPCS Implementation.

| Element | Ideal IPCS | Current Situation in SMPCS | Improvement Needed |
| --- | --- | --- | --- |
| Multidisciplinary teams | Fully integrated interdisciplinary teams | Present and functioning | Maintain and strengthen |
| Continuity of care | Seamless transitions across services | Partial continuity | Improve coordination across settings |
| Early patient identification | Early referral and integration | Often late referrals | Improve provider education |
| System integration | Unified service network | Fragmented services | Develop coordinated network |
| Community-based care | Strong outpatient and community PC | Limited availability | Expand rural services |
| Funding and resources | Stable funding and staffing | Limited funding | Increase resources |
| Information systems | Shared electronic care plans | Limited integration | Improve data sharing |
| Bereavement support | Integrated bereavement services | Limited support | Expand bereavement programs |

All participants agreed that SMPCS currently includes **multidisciplinary teams of palliative care specialists**, including nurses, physicians, nurse practitioners, physician assistants, social workers, dietitians, physiotherapists, pharmacists, chaplains, and professionals providing integrative therapies such as music, energy, and massage therapy.

Several participants reported **effective team coordination**, supported by regular interdisciplinary meetings and collaboration between inpatient palliative care teams and hospice services. Participants also emphasized the importance of **comprehensive, individualized care plans** that consider the needs of both patients and their families.

Some participants noted that **nurse-led case management** and **professional training in palliative care communication** were important strengths of the current system.

Despite these strengths, participants also identified several areas where improvements are needed. A commonly mentioned concern was the **lack of a unified palliative care network** connecting services across healthcare organizations and community settings. Participants described fragmentation in communication and coordination between services, which can lead to underutilization of palliative care resources. Participants also highlighted the need for improved **information sharing** systems to facilitate communication and coordination across services.

Issues related to **continuity of care** were also frequently reported. Participants described gaps between inpatient and outpatient care, as well as limited service availability outside regular hours, which may result in fragmented care for patients.

**Funding limitations and insurance coverage** were also identified as barriers affecting access to PCS. Participants indicated that limited resources and staffing shortages can make it difficult to provide adequate services across large rural geographic areas.

Participants also highlighted the need for **expanded outpatient and community-based PCS**, particularly for patients living in remote rural areas. Transportation challenges and geographic distance were identified as significant barriers to access.

Several interviewees emphasized the need for **improved education and training** for healthcare professionals, as well as greater education for families regarding the role and benefits of PCS. Participants also indicated that improving early identification of patients who could benefit from PCS remains an important area for development within the current system.In addition, participants noted the need for expanded **pediatric PCS** within the region.

Finally, participants identified opportunities to strengthen **collaboration between organizations**, develop **standardized models for palliative care delivery**, and enhance **bereavement support services** for families.

### 3.3 Conceptual Framework for Integrated Palliative Care in Rural Settings

Based on the themes identified in this study, a conceptual framework illustrating the key components of an integrated palliative care system in rural settings was developed (Figure 1). The framework synthesizes the findings from the interviews and illustrates how the identified elements operate across different levels of the healthcare system.

**Figure 1.**
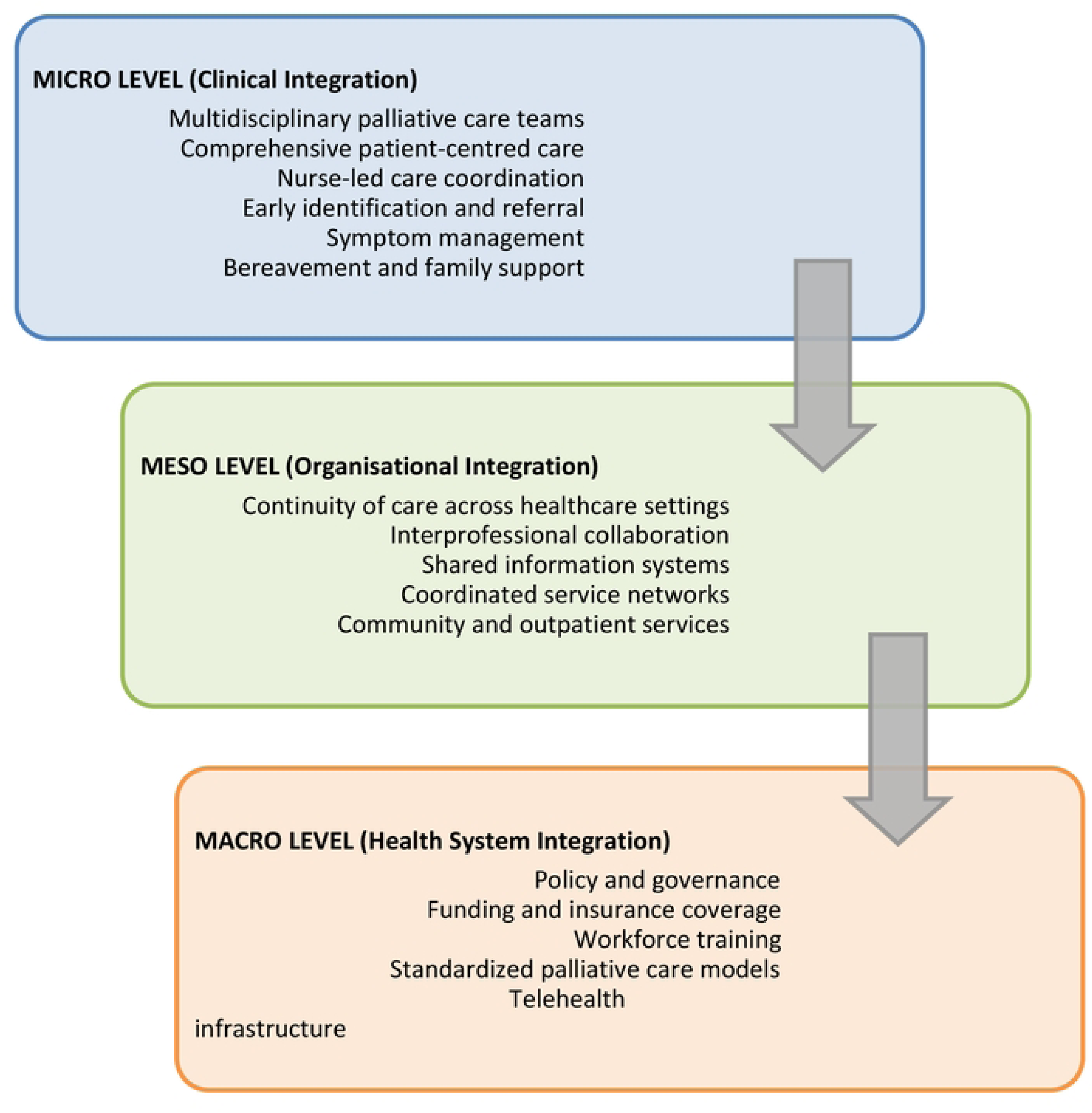
Conceptual Framework for Integrated Palliative Care in Rural Healthcare Systems.

The conceptual framework organizes the components of integrated palliative care into three interconnected levels: **micro (clinical), meso (organisational), and macro (health system)** levels. At the micro level, integration occurs primarily through multidisciplinary care teams, comprehensive patient-centered care, nurse-led care coordination, early identification of patients, and symptom management. These elements reflect the direct delivery of care to patients and families.

At the meso level, integration involves organisational mechanisms that support coordinated service delivery, including continuity of care across healthcare settings, interprofessional collaboration, shared information systems, and coordinated service networks linking hospital, community, and outpatient services.

Finally, at the macro level, system-level factors such as healthcare policies, funding mechanisms, insurance coverage, and the development of standardized models for palliative care delivery influence the broader implementation and sustainability of integrated PCS in rural healthcare systems.

## 4. DISCUSSION

This study explored the perspectives of healthcare professionals involved in the SMPCS regarding the level of integration of their PCS. Understanding these perspectives is particularly important in rural settings, where access to specialized services is often limited and coordination between services may present additional challenges. The findings of this study identified key components necessary for an ideal IPCS and highlighted both strengths and areas requiring improvement within the SMPCS to enhance system integration. This study contributes to the growing literature on integrated palliative care and health system integration by providing empirical evidence on how integration is experienced and operationalized within a rural healthcare system in the United States.

### 4.1 Essential Elements for an Ideal Integrated Palliative Care System

Participants identified several essential components for an ideal IPCS, including comprehensive patient-centered care, professional training and education, coordinated care networks, shared information systems, standardized frameworks and guidelines, interdisciplinary collaboration, continuity of care, case management, early patient identification, the use of technology to reach rural patients, and supportive community services and policies.

These elements are widely described in the literature on integrated care and palliative care systems (20–26). Their recurrence across diverse healthcare contexts highlights their fundamental importance for achieving integrated palliative care delivery.

Beyond their theoretical relevance, these components represent the practical foundation of effective PCS. Their identification reinforces the importance of coordinated service structures capable of addressing the complex needs of patients with chronic or life-limiting illnesses.

The need to strengthen IPCS is particularly important in rural settings, where structural barriers frequently limit access to specialized care. Rural populations often experience greater health disparities, geographic isolation, insufficient healthcare professionals in rural regions, and limited availability of PCS, all of which contribute to inequalities in palliative and end-of-life care delivery (27–29). These disparities highlight the importance of developing coordinated and integrated PCS that are specifically adapted to the needs of rural populations.

### 4.2 Elements Included or Requiring Improvement in the Southern Minnesota Palliative Care System

Participants reported that SMPCS currently incorporates several important components of IPCS, including multidisciplinary teams that are well-trained, coordinated care delivery, nurse-led case management, and the provision of comprehensive and individualized care. In addition, the use of virtual and face-to-face visits enables professionals to reach patients living in geographically dispersed rural areas. These findings are consistent with previous research highlighting the importance of interdisciplinary teams, coordinated service delivery, and professional training in improving care for patients with chronic and complex health conditions (30,31). The presence of these elements within SMPCS suggests that certain aspects of integration are already well established within the system.

However, participants also identified several areas requiring further development to strengthen integration. One of the most frequently mentioned aspects was the need to establish stronger palliative care networks and improve interorganizational collaboration. Integrated care networks and coordinated care pathways can significantly enhance communication between services and improve the distribution of clinical responsibilities across organizations involved in patient care (22). Strengthening these networks may help reduce care fragmentation and improve continuity of care between hospital, primary care, and community-based services (24).

Funding mechanisms, insurance coverage, and standardized protocols were also identified as areas requiring improvement. Standardized clinical protocols can help ensure consistent care delivery across healthcare settings and improve overall service quality. In addition, national strategies addressing funding, infrastructure, and resource allocation are essential to support integrated palliative care efforts (30).

In addition, recent policy discussions and proposed reductions in federal healthcare funding in the United States raise concerns regarding the sustainability of services that rely heavily on public reimbursement mechanisms such as Medicaid. Rural healthcare systems are particularly vulnerable to funding reductions because many facilities operate with limited financial margins and depend on public insurance programs to maintain essential services (32,33). Recent analyses suggest that reductions in federal health spending and Medicaid funding could place many rural hospitals at risk of service reductions or closure, further limiting access to healthcare for rural populations (34). These financial pressures may significantly affect the availability and expansion of PCS in underserved areas, where access to specialized services is already limited (35).

Participants also reported limitations related to reimbursement mechanisms for PCS. Insufficient reimbursement may restrict service availability and limit the number of visits that providers can offer to patients. These barriers can hinder effective management of chronic and life-limiting conditions and negatively impact the quality of care delivery. As highlighted in the WHO report on integrated care (36), financial policies that support coordinated care between health and social services are essential for improving access and sustainability of integrated care systems.

Early identification of patients requiring palliative care was also identified as an important area for improvement. Early integration of PCS has been associated with improved patient outcomes and better symptom management (37). Therefore, implementing structured referral systems and clinical screening tools may facilitate earlier identification of patients who could benefit from PCS (38).

Participants further emphasized the potential of telehealth and virtual care models to address geographical barriers in rural communities. Telemedicine can improve access to PCS for patients who face transportation challenges or limited availability of local providers (37,39). However, the effectiveness of telehealth interventions in rural settings may be constrained by limited broadband infrastructure and technological access(40).

Finally, participants emphasized the importance of strengthening community-based support systems for patients receiving palliative care. Community support networks, including family caregivers, social services, and volunteer organizations, can play a critical role in addressing care needs and supporting patients throughout the disease trajectory (26). However, research suggests that reliance on informal care networks in rural communities may vary significantly depending on family structure and available social support(41). These findings highlight the importance of integrating community-based resources into palliative care delivery systems.

Importantly, although this study was conducted in a specific rural region of the United States, the findings provide transferable insights into integrated palliative care delivery that may be applicable to rural healthcare systems internationally.

### 4.3 Integration of Care in the Southern Minnesota Palliative Care System

Integrated care is widely recognized as a multidimensional concept that operates across different levels of healthcare systems. According to Valentijn and colleagues(42), integrated care should be considered across three interrelated levels: micro (clinical care), meso (organizational and professional integration), and macro (system-level policies and governance).

The conceptual framework developed from the findings of this study (Figure 1) illustrates how these elements operate across the micro (clinical), meso (organisational), and macro (system) levels of integrated palliative care.

The findings of this study suggest that SMPCS demonstrates several facilitating elements at the micro level, particularly in relation to the delivery of PCS by multidisciplinary clinical teams. Micro-level integration is commonly observed in many healthcare systems, where coordinated clinical care models are implemented at the patient level (43). However, as noted in previous research, achieving sustainable integrated care requires coordination across all system levels. While clinical-level interventions may improve patient outcomes, they are often insufficient to achieve system-wide integration without supportive organizational structures and policy frameworks (43).

Consistent with these observations, the areas requiring improvement identified in this study were primarily located at the meso and macro levels. At the meso level, participants highlighted the need for stronger collaboration between organizations and improved continuity of care across healthcare settings. At the macro level, improvements in funding mechanisms, policy development, insurance coverage, and the establishment of unified palliative care networks were identified as necessary to support broader system integration.

Despite increasing recognition of the importance of PCS in the United States, access to these services remains uneven across regions (24). Even with the implementation of the Medicare Hospice Benefit, further policy and regulatory developments are needed to improve the accessibility and sustainability of PCS (44).

Overall, the findings of this study suggest that SMPCS demonstrates several elements of integrated care at the clinical level, while further development is needed at the organizational and system levels to achieve more comprehensive integration. Strengthening coordination between services, improving funding mechanisms, and developing unified care networks may support the continued evolution of integrated PCS in rural healthcare systems.

## 5. LIMITATIONS

This study provides insights into the integration of PCS in rural areas of southern Minnesota; however, several limitations should be considered when interpreting the findings. First, the limited availability of PCS in these rural settings restricted the recruitment of healthcare professionals eligible to participate in the study. As a result, the relatively small sample size may have limited the diversity of perspectives captured and the breadth of experiences represented.

Second, the qualitative design of this study aimed to explore in depth the experiences and perceptions of healthcare professionals rather than to produce statistically generalizable findings. Therefore, the results should be interpreted as context-specific insights that may not be directly generalizable to all healthcare systems or geographic regions; however, the findings offer transferable insights for similar rural healthcare contexts.

Finally, this study focused exclusively on the perspectives of healthcare professionals involved in PCS. Future research should incorporate the perspectives of patients, family caregivers, and community stakeholders in rural areas in order to provide a more comprehensive understanding of integrated palliative care experiences and needs.

Despite these limitations, the findings of this study offer several important implications for practice and policy. At the clinical level, strengthening multidisciplinary collaboration, early patient identification, and coordinated care pathways may enhance the delivery of PCS in rural communities. At the organisational level, improving communication systems, interorganizational collaboration, and care continuity across healthcare settings may support greater service integration.

At the system level, policymakers should consider strategies to strengthen funding mechanisms, expand insurance coverage, and support workforce development for palliative care in rural regions. In addition, investments in telehealth infrastructure and community-based services may help reduce geographic barriers and improve access to palliative care for underserved populations. Addressing these challenges across micro, meso, and macro levels of the healthcare system may support more sustainable and equitable palliative care delivery in rural communities.

## 6. CONCLUSIONS

PCS have demonstrated significant benefits for individuals living with chronic and life-threatening illnesses. Although PCS have expanded substantially in urban areas, rural communities remain underserved, creating important disparities in access to care. This study examined the integration of PCS within SMPCS and identified several elements that facilitate integrated care in this rural context.

The findings highlight key strengths within SMPCS, including well-trained and coordinated multidisciplinary teams that provide comprehensive and individualized care. Participants also emphasized the importance of nurse-led care management, patient education, and the use of both virtual and in-person visits to reach patients in geographically dispersed rural areas.

At the same time, several areas requiring improvement were identified. These include strengthening continuity of care, improving early identification of patients who may benefit from palliative care, expanding funding mechanisms and insurance coverage, increasing the availability of community-based and outpatient PCS, and developing unified networks that better integrate services across healthcare organizations.

Overall, the findings suggest that SMPCS demonstrates elements of integrated care at the clinical level, while additional efforts are needed to strengthen integration at the organizational and system levels. Addressing these challenges will be particularly important as the population of older adults continues to grow in the United States. Strengthening IPCS in rural settings may help reduce disparities in access to care and support more coordinated, equitable, and sustainable palliative care delivery, and these findings may also inform the development of integrated palliative care models in rural healthcare systems beyond the study setting.

## Data Availability

Data cannot be shared publicly because of the qualitative nature of the data and potential participant identification. Data are available from the Institutional Data Access / Ethics Committee (contact via authors by email) for researchers who meet the criteria for access to confidential data

## DECLARATIONS

### Ethics approval and consent to participate

This study was conducted in accordance with the ethical principles of the Declaration of Helsinki and applicable regulations for research involving human participants. The study was approved by the Institutional Review Board at Minnesota State University, Mankato (IRB#1877595; approved 5/19/2022).

### Consent to participates

All participants were informed about the study’s objectives, procedures, and potential risks and benefits, and they provided informed consent before participation. Participants were informed of their right to withdraw from the study at any time without consequence. Their personal information was kept confidential and used solely for the purposes of this study.

### Availability of data and materials

The datasets generated and/or analysed during the current study are not publicly available due to the qualitative nature of the data and potential participant identification but are available from the corresponding author on reasonable request.

### Authoŕs contribution

MMP, EV and KAA contributed to the conceptualization of the article. They all participated in data collection; MMP and KAA contributed to data curation and formal analysis. The three of them were involved in writing, reviewing and editing the Manuscript. MMP was responsible for the supervision. All authors have read and agreed to the final version of the manuscript.

## Acknowledgements

Not applicable.

## Author’s information

MMP is a Professor at the Department of Community Health and Life Cicle interventions, University of Vic-Central University (UVIC-UCC) of Spain. A member of the research group on Methodology, Methods, Models of Health and Social Outcomes (M3O). She obtained a bachelor’s degree in education at Blanquerna University Ramon Llull, Spain and a master’s degree in Comparative International Education at the University of Minnesota, USA. She graduated from St. Catherine University, USA, and obtained a post-baccalaureate degree in Nursing, later she completed a PhD in Comprehensive Care and Health services program at UVIC-UCC. She has more than 15 years of teaching experience at the university and the community level, and 5 years of experience as an oncology nurse. Her research is in the area of integrated palliative care and the chronically ill patient.

KAA is Dean and Professor in the College of Nursing at the University of Wisconsin Eau Claire, USA. She earned her diploma in nursing (Portland, OR, USA), Bachelor of Science in human development (2007) in Portland, Oregon, a master’s degree (2009) and Ph.D. (2015) in Nursing from the University of Wisconsin-Madison. Her Ph.D. dissertation entailed development of a sexual concerns questionnaire for women with gynecological cancer. Her current research focuses on individuals living with Alzheimer’s disease or related dementias, hospice and palliative care. She has extensive clinical practice and research experience in Alzheimer’s disease/associated dementias, and women’s health across the lifespan. Along with experience as a hospice RN and volunteer, she is also a trained End-of-Life doula as well an an End-of-Life Nursing Education Consortium (ELNEC) educator in core and geriatric curricula.

EV is an Associate Professor of Nursing at Minnesota State University, Mankato, with 15 years in nursing education and over 30 years as a registered nurse. She has 23 years of experience as an Advanced Practice Nurse Practitioner, specializing in adult and geriatric care, palliative and hospice services, and complex wound and continence management. She holds certifications in adult care, evidence-based practice mentoring, and higher education teaching. EV earned her DNP from Minnesota State University and her MSN from the University of Southern Maine. Her research focuses on improving palliative care in rural and underserved areas, and advancing excellence in online education and interprofessional leadership.

